# A two-arm, randomized, controlled, multi-centric, open-label Phase-2 study to evaluate the efficacy and safety of Itolizumab in moderate to severe ARDS patients due to COVID-19

**DOI:** 10.1101/2020.12.01.20239574

**Authors:** Suresh Kumar, Rosemarie de Souza, Milind Nadkar, Randeep Guleria, Anjan Trikha, Shashank R. Joshi, Subramanian Loganathan, Sivakumar Vaidyanathan, Ashwani Marwah, Sandeep N. Athalye

## Abstract

An uncontrolled increase in cytokine production may lead to systemic hyperinflammation, vascular hypo-responsiveness, increased endothelial permeability, hypercoagulation, multi-organ dysfunction and eventually death in moderate to severely ill COVID-19 patients. Targeting T-cells, an important driver of the hyperinflammatory response, in the treatment of COVID-19, could potentially reduce mortality and improve survival rates. Itolizumab is an anti-CD6 humanized monoclonal antibody with an immunomodulating action on T_effector_ cells that downregulates T-cell activation, proliferation and subsequent production of various chemokines and cytokines. The efficacy and safety of Itolizumab for the treatment of cytokine release syndrome in patients with moderate to severe acute respiratory distress syndrome (ARDS) due to COVID-19 was evaluated in a multi-centric, open-label, two-arm, controlled, randomized, phase 2 study. Eligible patients were randomized (2:1) to arm A (best supportive care + Itolizumab) and arm B (best supportive care). The primary outcome of interest was reduction in all-cause mortality 30 days after enrolment. Thirty-six patients were screened, 5 were treated as first dose sentinels and the rest were randomized, whilst 4 patients were considered screen failures. Two patients in the Itolizumab treatment arm discontinued prior to receiving the first dose and were replaced. At the end of 1 month, there were 3 deaths in arm B, and none in arm A (p= 0.0296). At the end of the follow-up period, more patients in Arm A had improved SpO2 without increasing FiO2 (p=0.0296), improved PaO2 (p=0.0296), and reduction in IL-6 (43 pg/ml vs 212 pg/ml; p=0.0296) and tumor necrotic factor-α (9 pg/ml vs 39 pg/ml; p=0.0253) levels. Itolizumab was generally safe and well tolerated, and transient lymphopenia (11 patients in Arm A) and infusion reactions (7 patients) were the commonly reported treatment related safety events. These encouraging results indicate that larger clinical trials are warranted to establish the role of Itolizumab in controlling immune hyperactivation in COVID-19.

## Introduction

Severe acute respiratory syndrome coronavirus-2 (SARS-CoV-2; COVID-19) poses a serious global concern for public health, with millions of people infected worldwide and more than a million dead [1,2]. Three escalating phases of SARS-CoV-2 disease progression have been reported [3,4]. During the early infection phase, the virus infiltrates the lung parenchyma and proliferates causing mild constitutional symptoms. The second phase is characterized by adaptive immunity stimulation (vasodilation, endothelial permeability, leukocyte recruitment and tissue damage) with lung injury and hypoxemia as underlying causes of the respiratory dysfunction (pulmonary phase). Lung vascular thrombosis may be predominant during this phase. In the third phase (hyperinflammation phase), systemic inflammatory response may set in leading to increased production of a series of cytokines and prime adaptive T- and B-cell responses [5]. SARS-CoV-2 infection sets off an inflammatory cascade resulting in an increased release of pro-inflammatory cytokines and chemokines, especially IL-1, IL-6, IL-12, IFN-γ, and TNF-α [6]. These proinflammatory molecules potentiate a Th1 (T helper-1) response, causing the recruitment of monocytes and T lymphocytes resulting in peripheral lymphopenia and higher neutrophil:lymphocyte ratio typically observed in patients suffering from COVID-19 [7–9]. If untreated, this cytokine release syndrome may lead to vascular hypo-responsiveness, increased endothelial permeability, hypercoagulation, multi-organ dysfunction and eventually death [4].

Targeting T cells and their involvement in cytokine release syndrome during the management of SARS-CoV-2 disease has been one of the therapeutic strategies adopted to improve survival rates and reduce mortality. Unlike the other anti-inflammatory agents such as Tocilizumab (IL-6 inhibitor), Sarilumab (IL-6R inhibitor), and Anakinra (IL-1R inhibitor), Itolizumab (CD6 inhibitor) has an upstream immunomodulating mechanism of action [10–12]. CD6 is a costimulatory receptor differentially expressed on T-cells, subsets of innate lymphoid and natural killer cells, but not on T regulatory cells [13–15]. It is implicated in the pathogenesis of multiple autoimmune and inflammatory diseases. The binding of CD6 to the activated leukocyte cell adhesion molecule (ALCAM), expressed in both the antigen presenting cells and endothelial/epithelial tissue, including the blood-brain barrier, skin, gut, lung and kidney, can modulate T-cell activity and trafficking [16].

Itolizumab is a humanized IgG1 kappa anti-CD6 monoclonal antibody that binds to domain 1 of human CD6. It selectively targets the CD6-ALCAM pathway resulting in decreased levels of IFN-γ, IL-6, and TNF-α through Th-1 pathway and IL-17, IL-6, TNFα through Th-17 pathway [17,18]. Itolizumab thereby leads to a reduction in the T-cell infiltration at the sites of inflammation, without inducing T-cell or B-cell depletion [17].

Itolizumab has been approved in India for the treatment of moderate-to-severe chronic plaque psoriasis for the last seven years [19,20] and in addition to a favorable safety profile in Phase 2 and Phase 3 trials, has shown promising results when used in psoriatic arthritis [10] and rheumatoid arthritis [21]. Itolizumab has demonstrated a durable therapeutic effect which is noted even after discontinuation of the treatment [22] in the management of psoriatic arthritis [23,24] and rheumatoid arthritis [21]. Previous studies have shown the impact of Itolizumab on human Th1 cells [17]. It has been demonstrated that even under the classical co-stimulation by anti-CD3 and anti-CD28 [25], Itolizumab is able to down-regulate the expression of key Th17 determining transcription factors and effector cytokines (i.e., IL-17) in addition to decreasing Th1 effector cytokine (IFN-γ) (Figure 1).

**Figure 1:**
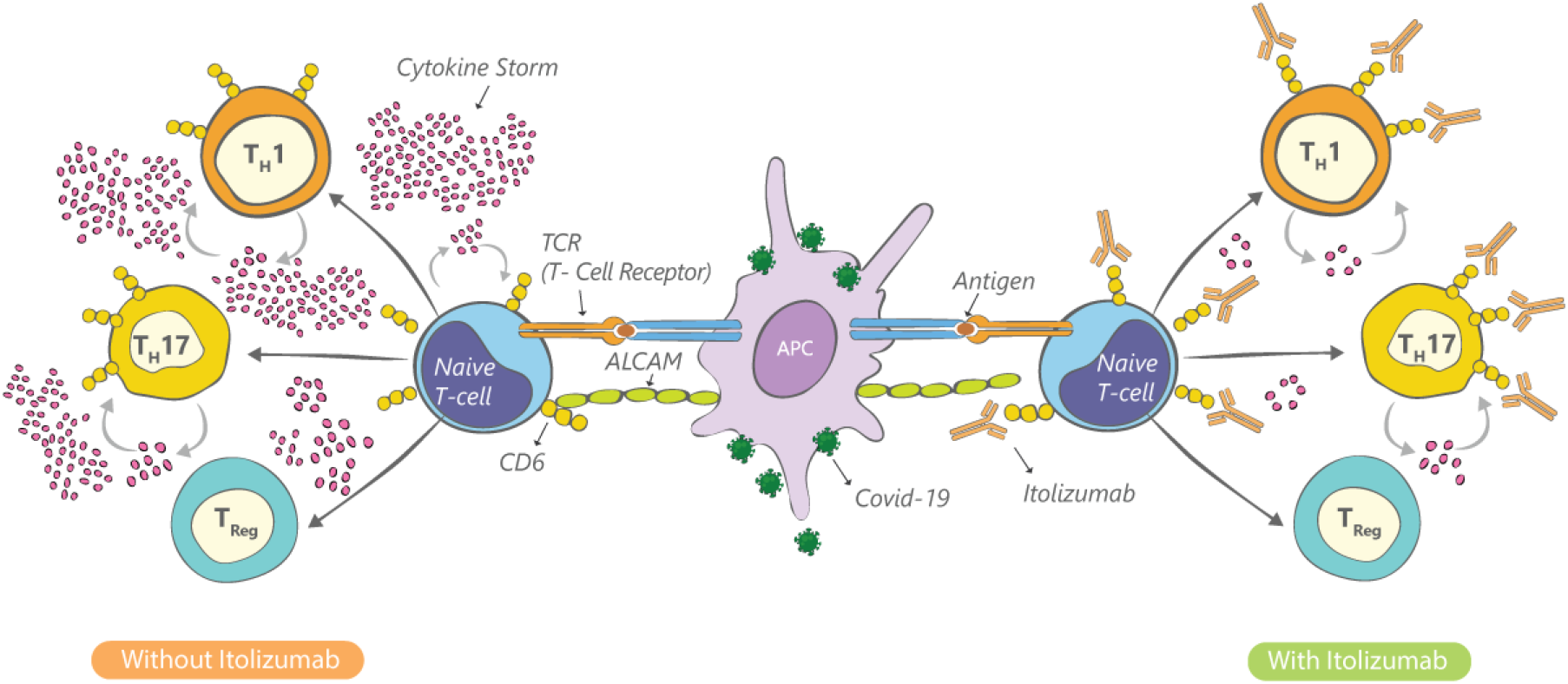
Itolizumab mechanism of action in COVID-19 infection.

CD6 is expressed mainly on effector T cells (Teff). CD6 stimulates ALCAM mediated T-cell activation and subsequently pro-inflammatory cytokines release. Itolizumab inhibits T-cell activation and lowers major pro-inflammatory cytokines of the Th1/Th17 pathways.

We hypothesized that Itolizumab will control the pro-inflammatory cytokine release in COVID-19 patients by immunomodulation of Teff function and trafficking to the inflammation site, sparing Tregs and preserving the anti-viral response, reducing morbidity and mortality. The current study was undertaken to estimate the efficacy and safety of Itolizumab in the treatment of cytokine release syndrome in patients with moderate to severe acute respiratory distress syndrome (ARDS) due to COVID-19.

## Methods

### Study design

This was an open-label, two-arm, randomized, controlled, multi-centric, phase-2 study conducted in 4 designated COVID-19 hospitals in India. Initial dosing was done for first five patients in a staggered manner wherein after a patient was dosed, safety was monitored for 24-48 hours prior to dosing of the next patient. Once all five patients were dosed in this staggered manner, subsequent patients were enrolled such that study had patients randomized in a 2:1 ratio. Randomization was centrally done using computer-generated sequences (SAS version 9.4). Patients who were randomized, but did not receive the full infusion, were considered unevaluable and the same randomization code was used for allocation of the next patient enrolled by the study site. The CONSORT flow diagram for the study is summarized in Figure 2. The study was initiated on May 2, 2020, and all patients were followed up for 30 days, with the study closing out on July 7, 2020 when the follow up period of the final patient was completed and all patients in the trial had either been discharged from clinical care or died from COVID-19 complications.

**Figure 2.**
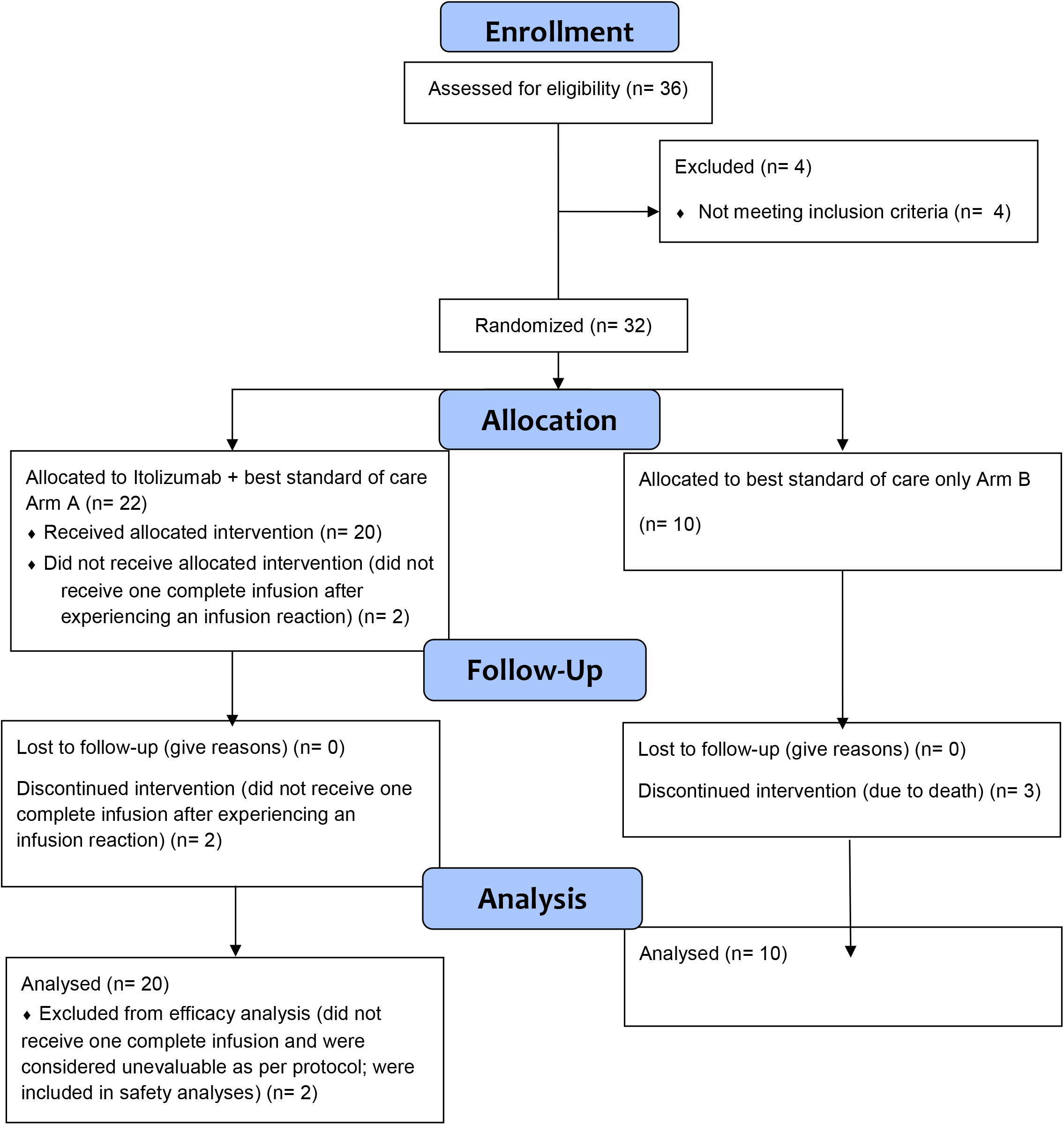
CONSORT 2010 Flow Diagram.

### Study subjects

Adult male or female patients above 18 years, who tested positive for virologic diagnosis of SARS-CoV-2 infection (RT-PCR), and who were hospitalized due to clinical worsening with oxygen saturation of ≤94% at rest in ambient air, were eligible for randomization if they had either moderate to severe ARDS or high levels of proinflammatory markers. Patients were defined to have moderate to severe ARDS if they had PaO2/Fio2 ratio of < 200 or more than 25% deterioration from the immediate previous value. Alternatively, the proinflammatory markers included were baseline serum ferritin level ≥ 400 ng/mL or IL-6 levels greater than 4 times of upper limits of normal value.

Major exclusion criteria included - known severe allergic reactions to monoclonal antibodies, an active tuberculosis (TB) infection/inadequately treated tuberculosis/latent tuberculosis, on oral anti-rejection or any immune-suppressive drugs in last 6 months, those who participated in other drug clinical trials using anti-IL-6 therapy. Patients with a known history of Hepatitis B, Hepatitis C or HIV, absolute neutrophils count (ANC) <1000 / mm^3^, platelet count <50,000 / mm^3^ and absolute lymphocyte count (ALC) <500/mm^3^ were also excluded.

### Study Settings

The study was carried out at four COVID-19 specific, tertiary hospitals in India. Two of these sites were in New Delhi, and two were in Mumbai. All four sites were tertiary, teaching hospitals, with considerable experience of undertaking clinical trials.

### Treatments

Most commonly used therapies as part of best supportive care in both treatment arms included oxygen, antibiotics, hydroxychloroquine, antivirals, steroids, low-molecular-weight heparin, and vitamin supplements.

The dose of Itolizumab was calculated and diluted in 250 ml of normal (0.9%) saline; this was allowed to reach room temperature prior to infusion. Itolizumab infusion in Arm A started after premedication with hydrocortisone 100 mg i.v (or equivalent short acting glucocorticoid) and Pheniramine 30 mg per i.v. about 30 ± 10 minutes prior to infusion. Patients were initiated on 1.6 mg/kg dose iv infusion of Itolizumab and continued with 0.8 mg/kg dose weekly regimen as required. Subsequent doses were modified, deferred, or stopped as per the investigator’s discretion if the patient recovered. The Itolizumab infusion was administered over a period not less than 120 minutes, using an infusion set with an in-line, sterile, non-pyrogenic, low protein-binding filter (pore size of 1.2 μm or less). Approximately 50 mL of infusion solution was administered during the first hour, followed by remaining solution (approximately 200 mL) in the next hour. Infusion period could be extended up to 8 hours for medical reasons, particularly if the patient experienced infusion related reactions, which needed medical attention prior to re-initiation of infusion. Itolizumab was not infused concomitantly in the same IV line with any other agents.

### Ethics

This study was carried out in accordance with the ethical principles described in the Declaration of Helsinki (64th WMA General Assembly, Fortaleza, Brazil, October 2013), the International Council for Harmonization Good Clinical Practice (ICH GCP) E6 (R2), and New Drugs and Clinical Trials Rule 2019 issued by the Government of India. The study received approvals from the IECs/IRBs of all the participating sites. All the IECs/IRBs had active CDSCO registration at the time of approving this study. Subjects provided written informed consent prior to initiation of the study procedures. Study data was periodically reviewed by a data and safety monitoring board (DSMB).

### Study Objectives and Endpoints

The primary objective of this study was to estimate the efficacy and safety of Itolizumab in the treatment of cytokine release syndrome in patients with moderate to severe ARDS due to COVID-19. Secondary Objective was to assess possible correlations/associations between cytokine markers and clinical efficacy/safety.

The study’s primary outcome measures included:

1. Reduction in mortality one month after randomization
2. Reduction in the proportion of patients with deteriorating lung functions, as measured by:
  a. Stable SpO2 without increasing FiO2
  b. Stable PaO2 without increasing FiO2
3. Reduction in proportion of patients who needed non-invasive ventilation, invasive mechanical ventilation/endotracheal intubation, and high flow nasal oxygen
4. Reduction in inflammatory markers: Ferritin, D-dimer, LDH, CRP.

Key secondary outcome measures included measurement of:

1. Biomarkers such as IL-6, TNF-α, IL-17A
2. Absolute lymphocyte count
3. PaO2/FiO2 ratio calculated from arterial blood gas analyses
4. Safety: Number of participants with treatment-related side effects as assessed by Common Terminology Criteria for Adverse Event (CTCAE) version 5.0

### Biomarker assessments

Blood samples were collected for analysis of cytokines/chemokines.

### Statistical analysis

For this Phase 2 study, we considered enrolling 30 patients. Continuous variables were summarized using descriptive statistics such as mean, standard deviation, 95% confidence interval (CI), or median with range, as appropriate. Categorical variables were summarized using proportions (counts and percentages). Comparisons between proportions was done using Fisher’s exact test since the sample size was small. For continuous variables, change from baseline or trend in change over time were tabulated, as appropriate. All statistical tests were performed at 5% level of significance (two-sided test) and p-value<0.05 considered statistically significant. All statistical analysis was performed using SAS^®^ (version 9.4) software.

### Trial Registration Details

The trial protocol was registered with the Clinical Trials Registry of India (CTRI). The CTRI registration number is CTRI/2020/05/024959 and it can be accessed at this link: http://ctri.nic.in/Clinicaltrials/showallp.php?mid1=42878&EncHid=&userName=itolizumab. The trial was prospectively registered with the CTRI, which is the government mandated registry for trials in India.

## Results

### Participant disposition and baseline characteristics

A total of 36 patients were screened of which 4 were considered screen failures; 1 patient was COVID-19 negative and ALC count of 3 patients was <500 cells/cu.mm. A total of 32 patients were randomized: 22 patients in Arm A and 10 patients in Arm B (Table 1). Two patients from Arm A discontinued treatment prior to completion of first dosing due to infusion related reactions and were replaced as defined above. The events of infusion related reactions (IRRs) resolved on the same day and the patients continued to receive best supportive care. A total of 27 patients (Arm A: 20 and Arm B: 7) completed the study; 3 patients in Arm B discontinued due to death. All 20 patients in Arm A had at least one complete infusion of Itolizumab; of these, 7 patients had two infusions; 3 patients had three infusions and 4 patients had four infusions.

**Table 1:** Participant disposition.

| Variable | Arm A<br>(N=20) | Arm B<br>(N=10) |
| --- | --- | --- |
| FAS Population,* n (%) | 20 (100.00) | 10 (100.00) |
| Safety Population,** n (%) | 22 (110.00) | 10 (100.00) |
| Completed the study, n (%) |  |  |
| Completed 30 days follow up in hospital | 4 (20.00) | 1 (10.00) |
| Early discharged** | 16 (80.00) | 6 (60.00) |
| Discontinued | - | 3 (30.00) |
| Reasons for Discontinuation, n (%) |  |  |
| Death | - | 3 (30.00) |

\*FAS: Full analysis set defined as all patients randomized and those who received at least one full dose of Itolizumab (Arm A); Safety population was defined as all patients randomized (in Arm B) and those who received partial or full dose of Itolizumab (in Arm A)
\*\*2 subjects could not complete even one dosing and were replaced as per protocol

The median age of patients in Arm A was 50.5 years and in Arm B was 49.5 years (Table 2). All patients were of Asian ethnicity with most of the patients being male (Arm A: 95% and Arm B: 70%).

**Table 2:**
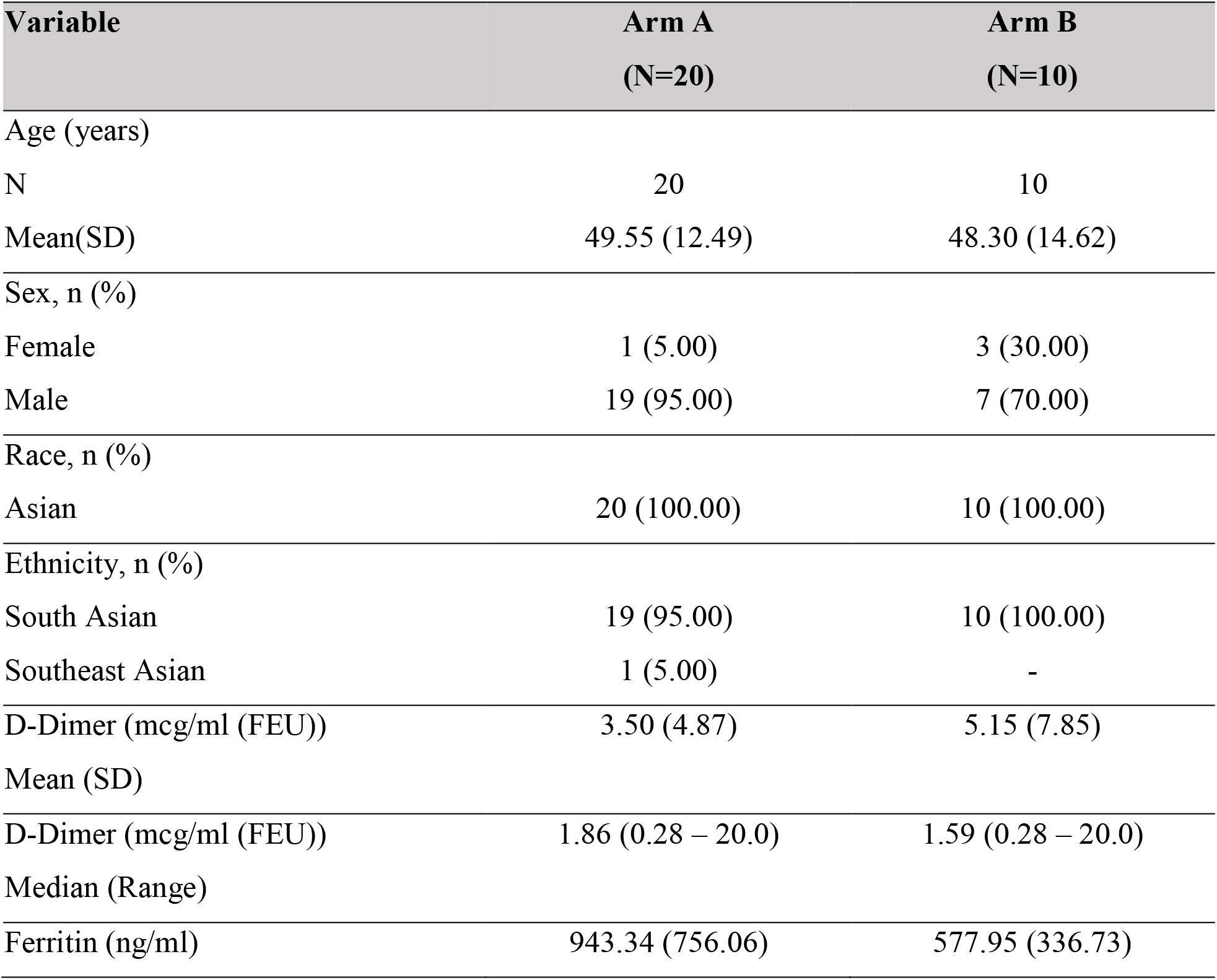
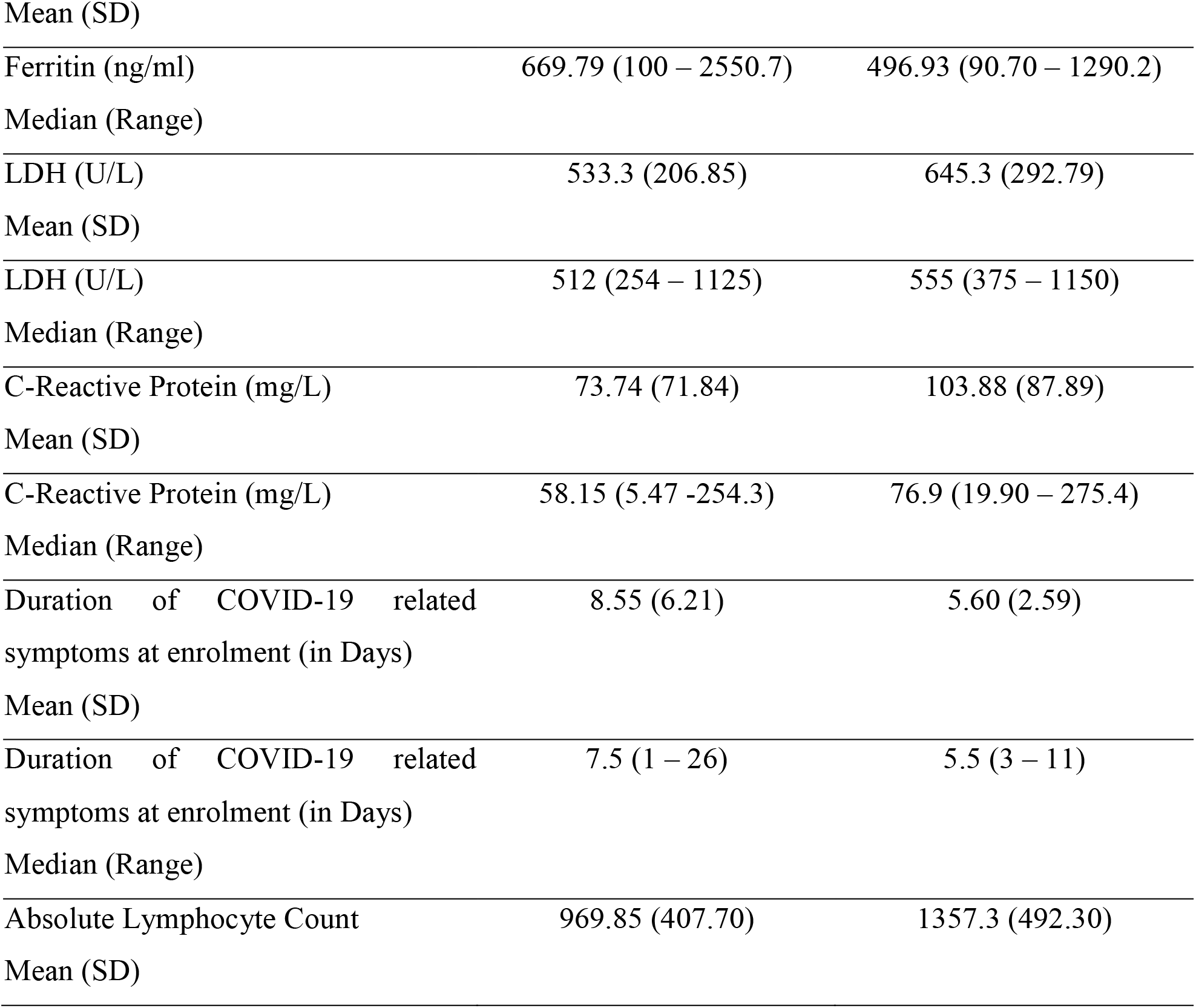
Demographic and baseline characteristics.

| **Variable** | **Arm A**<br>**(N=20)** | **Arm B**<br>**(N=10)** |
| --- | --- | --- |
| Age (years) |  |  |
| N | 20 | 10 |
| Mean(SD) | 49.55 (12.49) | 48.30 (14.62) |
| Sex, n (%) |  |  |
| Female | 1 (5.00) | 3 (30.00) |
| Male | 19 (95.00) | 7 (70.00) |
| Race, n (%) |  |  |
| Asian | 20 (100.00) | 10 (100.00) |
| Ethnicity, n (%) |  |  |
| South Asian | 19 (95.00) | 10 (100.00) |
| Southeast Asian | 1 (5.00) | - |
| D-Dimer (mcg/ml (FEU)) | 3.50 (4.87) | 5.15 (7.85) |
| Mean (SD) |  |  |
| D-Dimer (mcg/ml (FEU)) | 1.86 (0.28 – 20.0) | 1.59 (0.28 – 20.0) |
| Median (Range) |  |  |
| Ferritin (ng/ml) | 943.34 (756.06) | 577.95 (336.73) |

| Variable | Arm A<br>(N=20) | Arm B<br>(N=10) |
| --- | --- | --- |
| Mean (SD) |  |  |
| Ferritin (ng/ml) | 669.79 (100 – 2550.7) | 496.93 (90.70 – 1290.2) |
| Median (Range) |  |  |
| LDH (U/L) | 533.3 (206.85) | 645.3 (292.79) |
| Mean (SD) |  |  |
| LDH (U/L) | 512 (254 – 1125) | 555 (375 – 1150) |
| Median (Range) |  |  |
| C-Reactive Protein (mg/L) | 73.74 (71.84) | 103.88 (87.89) |
| Mean (SD) |  |  |
| C-Reactive Protein (mg/L) | 58.15 (5.47 -254.3) | 76.9 (19.90 – 275.4) |
| Median (Range) |  |  |
| Duration of COVID-19 related symptoms at enrolment (in Days) | 8.55 (6.21) | 5.60 (2.59) |
| Mean (SD) |  |  |
| Duration of COVID-19 related symptoms at enrolment (in Days) | 7.5 (1 – 26) | 5.5 (3 – 11) |
| Median (Range) |  |  |
| Absolute Lymphocyte Count | 969.85 (407.70) | 1357.3 (492.30) |
| Mean (SD) |  |  |

The mean duration of COVID-19 related symptoms at enrolment was 8.6 days and 5.6 days for Arms A and B, respectively, and the difference was not statistically significant. The most frequently reported COVID-19 related symptoms in both treatment arms were fever and dyspnea, followed by cough and tachypnea. Hypertension was the most common active co-morbid condition (20% in each arm).

### Primary outcome measures

#### 1. Mortality at 1 month

Itolizumab treatment had a noticeable improvement on patient’s survival through reduction in 1-month mortality rate. A statistically significant difference (p=0.0296; 95% CI= -0.3 [-0.61, -0.08])) in the 1-month mortality rate was observed between the 2 treatment arms. Three deaths were reported in Arm B on Days 4, 5 and 12 (1 due to acute respiratory distress syndrome and 2 due to respiratory failure). There were no deaths in Arm A.

#### 2. Lung function determined by SpO2 and PaO2

##### a. Stable/improved SpO2

A higher proportion of patients in Arm A had stable/improved SpO2 without increasing FiO2 in all the post-baseline assessment visits in comparison to Arm B (Table 3). A significant difference was observed between the 2 arms from Day 21 onwards; 100% of the participants in Arm A showed favorable outcomes compared to only 70% in Arm B (p=0.0296).

**Table 3:** Patients with Stable/Improved SpO2 without Increasing FiO2*.

| Visit | Arm A<br>(N=20) | Arm B<br>(N=10) | P-value |
| --- | --- | --- | --- |
| Day 7 | 17 (85.00) | 5 (50.00) | 0.0778 |
| Day 14** | 19 (95.00) | 7 (70.00) | 0.0952 |
| Day 21 | 20 (100.00) | 7 (70.00) | 0.0296 |
| Day 30 | 20 (100.00) | 7 (70.00) | 0.0296 |

\* Stable SpO2 was defined as absence of increase in FiO2 to maintain Spo2 ≥ 92% and improvement of SpO2 was defined as decrease in FiO2 to maintain SpO2 $>$ 92%.
\*\* Patients improved/ weaned off O2, the observation was carried forward; 3 patients in Arm B died on Day 4, 5 and 12; p-value between arm is estimated using Fisher's exact test (p-value $<$ 0.05 is considered significant)

##### b. Stable/Improved PaO2

A higher proportion of patients in Arm A had stable PaO2 without increasing FiO2 in all the post-baseline assessment visits in comparison to Arm B (Table 4). Significant difference was observed Day 21 onwards; 100% in Arm A compared to 70% in Arm B (p=0.0296).

**Table 4:** Patients with Stable/Improved PaO2 Without Increasing FiO2*.

| Visit | Arm A<br>(N=20) | Arm B<br>(N=10) | P-value |
| --- | --- | --- | --- |
| Day 7 | 18 (90.00) | 6 (60.00) | 0.1413 |
| Day 14** | 19 (95.00) | 7 (70.00) | 0.0952 |
| Day 21 | 20 (100.00) | 7 (70.00) | 0.0296 |
| Day 30 | 20 (100.00) | 7 (70.00) | 0.0296 |

\* Stable PaO<sub>2</sub> was defined as up to 10% change in PaO<sub>2</sub>/FiO<sub>2</sub> ratio from baseline while an improvement of PaO<sub>2</sub> was defined as > 10% improvement in PaO<sub>2</sub>/FiO<sub>2</sub> ratio from baseline (including patients weaned off oxygen).
\*\* Patients improved/ weaned off O<sub>2</sub>, the observation was carried forward; 3 patients in Arm B died on Days 4, 5 and 12; p-value between arm is estimated using Fisher's exact test (p-value <0.05 is considered significant)

#### 3. Non-invasive ventilation, invasive mechanical ventilation/endotracheal intubation, and high flow nasal oxygen

In Arm A, there were 5 patients on NIV (BiPAP or CPAP) at baseline that improved and came off NIV by Day 14. In Arm B, there were 4 patients on NIV at baseline of which 1 patient improved and came off NIV by Day 14. Condition of the 3 remaining patients in Arm B, which included 1 patient who continued to be on NIV and 2 patients who progressed to IMV before Day 7, further worsened and all of them died by Day 12. All the patients in Arm A were progressively weaned off oxygen by Day 30, with 5, 14, 18 and 20 patients getting weaned by days 7, 14, 21 and 30, respectively. In Arm B, 2, 4, 6 and 7 patients were progressively weaned off oxygen on Days 7, 14, 21 and 30 respectively.

#### 4. Inflammatory Markers (related to primary outcomes)

##### a. Ferritin

Baseline ferritin was high in Arm A compared to Arm B (943.34 ng/mL vs 577.95 ng/mL). In Arm A, the mean ferritin reduced to 303.50 (SD 210.93) ng/dL and 189.22 (SD 129.96) ng/dL on Day 14 and 30, respectively. In Arm B, the ferritin was 367.68 (SD 130.22) ng/dL and 285.25 (SD 157.76) ng/dL on Days 14 and 30, respectively. A greater reduction from baseline in serum ferritin levels was seen in Arm A (−479.3 (620.95) ng/dL) in comparison to Arm B (−234.4 (405.67) ng/dL) at day 30.

##### b. D-dimer

Baseline D-dimer was higher in Arm B (5.15 (SD 7.85) µg/mL) compared to Arm A (3.50 (SD 4.87) µg/mL). In Arm A, mean D-dimer reduced to 2.83 (SD 5.46) µg/mL and 0.41 (SD 0.18) µg/mL on Days 14 and 30, respectively. In Arm B, mean D-dimer was 0.86 (SD 0.71) µg/mL and 1.15 (SD 0.37) µg/mL on Days 14 and 30, respectively. Eight patients in Arm A and five patients in Arm B received low-molecular-weight heparin.

##### c. LDH

Baseline LDH was comparable in both arms; 533.30 (SD 206.85) U/L in Arm A and 645.30 (SD 292.79) U/L in Arm B. In Arm A, mean LDH reduced to 381.47 (SD 181.45) U/L and 208.67 (SD 40.72) U/L on Days 14 and 30, respectively. In Arm B, mean LDH was 330.20 (SD 91.63) U/L and 456.50(SD 173.24) U/L on Days 14 and 30, respectively.

##### d. CRP

Baseline CRP was numerically higher in Arm B; 73.74 (SD 71.84) mg/L in Arm A vs 103.88 (SD 87.89) mg/L in Arm B. After randomization in Arm A, mean CRP reduced to 6.45 (SD 4.14) mg/L and 13.69 (SD 20.45) mg/L on Days 14 and 30, respectively. In Arm B, mean CRP was 14.36 (9.29) mg/L and 3.05 (2.62) mg/L on Days 14 and 30, respectively.

The improvement in the biomarker status over time is captured in table 5 which outlines the mean change from baseline values over time. Number of patients at each time point varied due to patients reaching the end of follow up either due to discharge from clinical care or death.

**Table 5.** Mean change from baseline values for inflammatory markers.

| **Ferritin (ng/mL)** |  |  |  |  |
| --- | --- | --- | --- | --- |
|  | **Day 7** | **Day 14** | **Day 21** | **Day 30** |
| Arm A | -117.8 | -713.9 | -780.9 | -479.3 |
| N* | 18 | 15 | 11 | 3 |
| Arm B | -87.05 | -209.6 | 4238 | -234.4 |
| N** | 7 | 5 | 3 | 2 |
| **D-dimer (µg/mL FEU)** |  |  |  |  |
|  | **Day 7** | **Day 14** | **Day 21** | **Day 30** |
| Arm A | -1.43 | -0.45 | -4.35 | -2.63 |
| N* | 18 | 12 | 11 | 3 |
| Arm B | 2.3 | -0.68 | 8.54 | -0.35 |
| N** | 7 | 4 | 2 | 2 |
| **LDH (U/L)** |  |  |  |  |
|  | **Day 7** | **Day 14** | **Day 21** | **Day 30** |
| Arm A | -134 | -195.8 | -308.1 | -212.7 |
| N* | 18 | 15 | 11 | 3 |
| Arm B | -44.29 | -195.2 | 155.33 | -97 |
| N** | 7 | 5 | 3 | 2 |
| **CRP (mg/L)** |  |  |  |  |
|  | **Day 7** | **Day 14** | **Day 21** | **Day 30** |
| Arm A | -61.69 | -81.65 | -90.99 | -103.2 |
| N* | 18 | 16 | 11 | 3 |
| Arm B | -103.6 | -107.2 | -127.5 | -127.6 |
| N** | 8 | 5 | 3 | 2 |

\* Number of patients in Arm A at given time points
\*\* Number of patients in Arm B at given time points

### Secondary outcome measures

#### 1. Biomarkers

##### a. IL-6

Mean baseline value of IL-6 was comparable in both arms; 159.09 pg/mL in Arm A and 162.16 pg/mL in Arm B. A significant decline (p=0.0269) in mean IL-6 levels post first infusion was seen in Arm A (42.98 pg/mL) compared to Arm B (211.52 pg/mL) (Figure 3).

**Figure 3.**
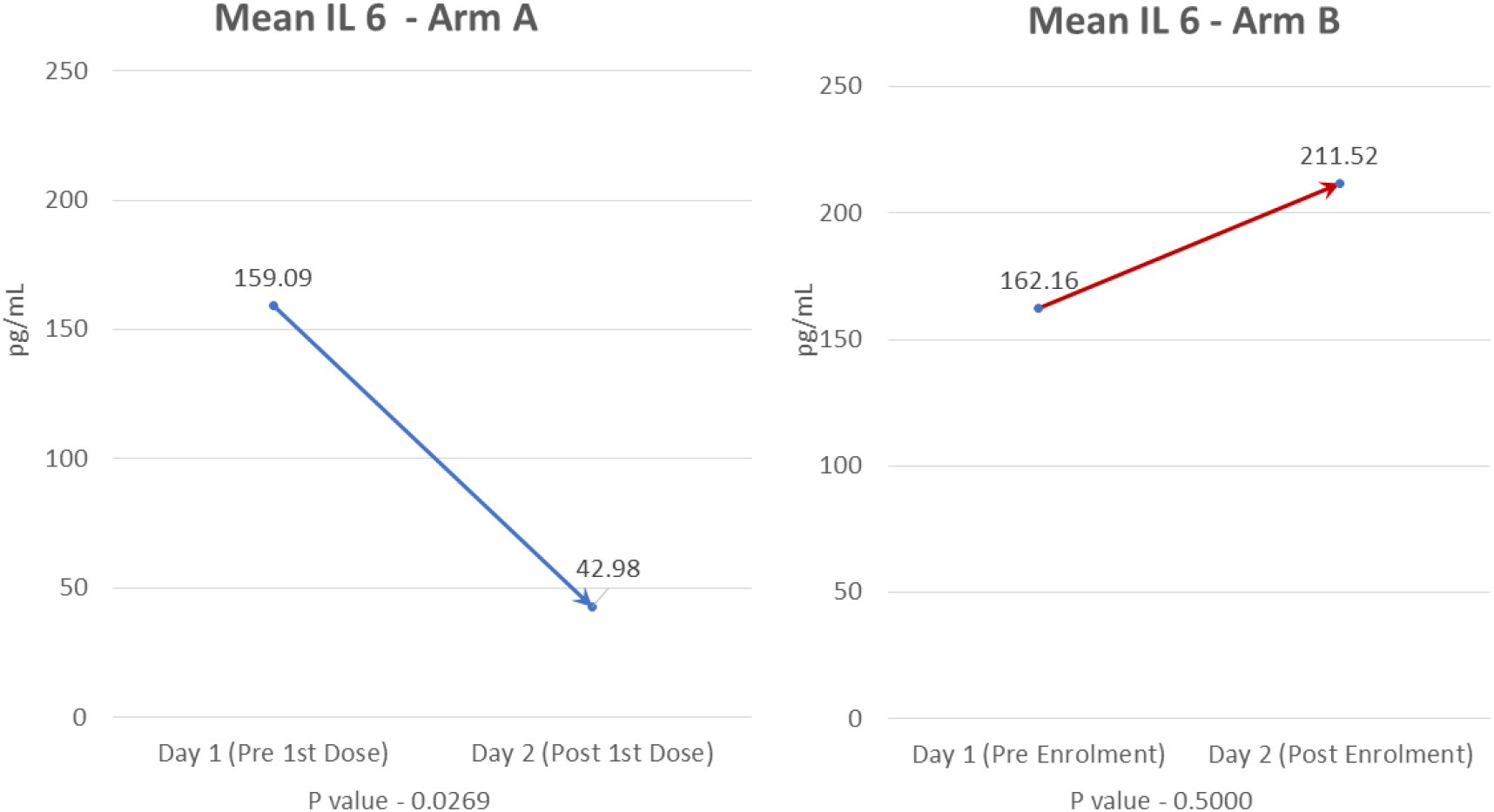
Mean IL-6 values.

##### b. TNF-α

Mean baseline value of TNF α was higher in Arm A (43.64 pg/mL) than in Arm B (11.26 pg/mL). A significant decline (p= 0.0253) in mean TNF-α levels post first infusion was seen in Arm A (8.87 pg/mL) compared to Arm B (39.19 pg/mL) (Figure 4).

**Figure 4:**
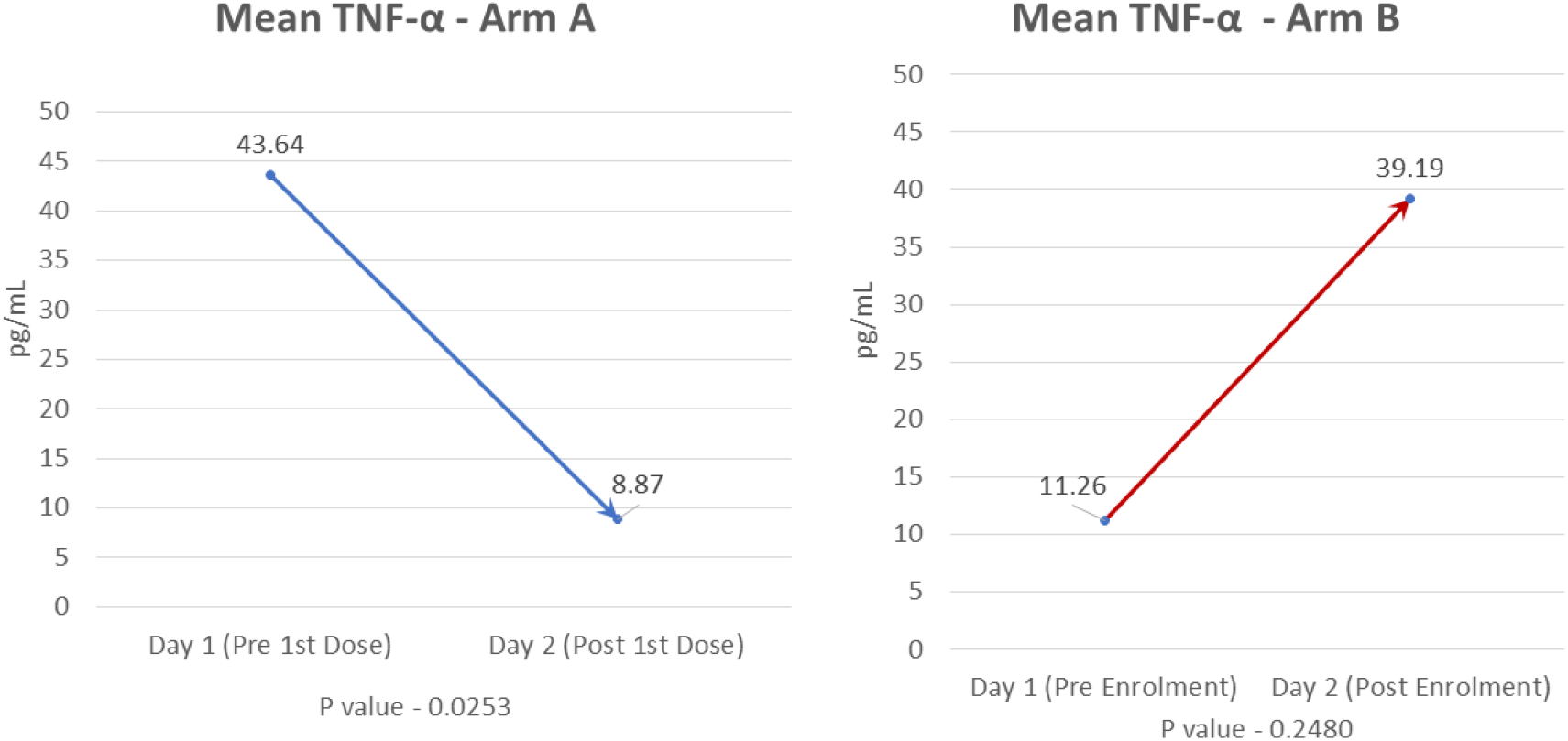
Mean TNF-α.

##### c. IL-17A

Mean baseline value of IL-17A was comparable in both arms; 10.36 pg/mL in Arm A and 9.83 pg/mL in Arm B. A notable decline in mean IL-17A levels post first infusion was seen in Arm A (6.75 pg/mL) unlike in Arm B, where there was an increase (14.75 pg/mL).

#### 2. Absolute Lymphocyte Count

Baseline ALC was numerically lower in Arm A (969.85 cells per mm^3^) than in Arm B (1357.3 cells per mm^3^). A gradual increase over time in mean ALC was seen in Arm A in comparison to Arm B (Figure 5). Eleven patients in arm A and 2 patients in Arm B had a grade 3 event of post-infusion lymphopenia, which was transient and recovered spontaneously by day 7.

**Figure 5:**
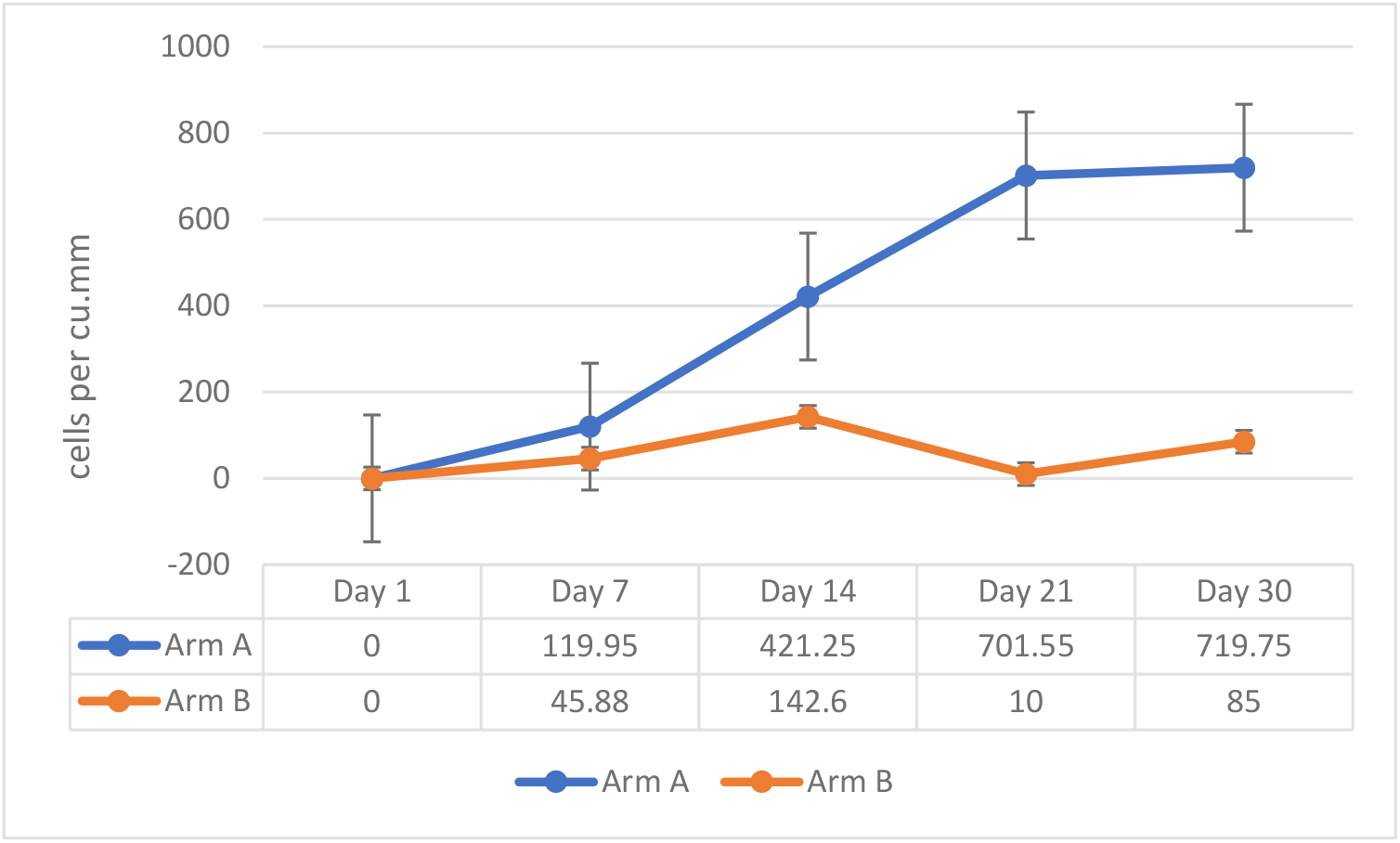
Mean Absolute Lymphocyte Count.

#### 3. PaO2 / FiO2 ratio

At Baseline, mean PaO2/FiO2 ratio was numerically higher in Arm A (126.57; SD 38.31) vs Arm B (114.05; SD 30.93). PaO2/FiO2 ratio gradually increased over time in Arm A in comparison to Arm B. The change from baseline observed at various time points is mentioned in Table 6. Given that there were censoring events (discharged from care or death) over time, the number of patients in each arm varied at each time point.

**Table 6.** Mean PaO2/FiO2 ratio over time.

|  | Baseline | Day 7 | Day 14 | Day 21 | Day 30/EOS |
| --- | --- | --- | --- | --- | --- |
| Arm A (n) | 20 | 16 | 14 | 8 | 3 |
| Mean (SD) | 126.57(38.31) | 203.50(95.51) | 283.43(104.26) | 350.25(70.36) | 397.67(15.63) |
| Arm B (n) | 10 | 6 | 5 | 3 | 0 |
| Mean (SD) | 114.05(30.93) | 184.53(95.51) | 338.40(42.57) | 398.33(24.01) |  |

#### 4. Safety

During the treatment period, a total of 22 patients experienced at least one treatment emergent adverse event (TEAE); 18 (81.82%) patients in Arm A and 4 (40.00%) patients in Arm B. Transient lymphocyte count decrease was the most commonly reported TEAE in both arms in addition to lower respiratory tract infection, ARDS and respiratory failure in Arm B (Table 7). Lymphocyte count decrease was the most frequently reported study drug related TEAE reported in 50% of patients in Arm A (n=11). These events were reported between Day 2 to Day 4 and returned to normal by Day 7.

**Table 7.** Treatment emergent adverse events by treatment group (Safety population)

| **System Organ Class** | **Arm A** | **Arm B** |
| --- | --- | --- |
| **Preferred Term** | **(N=22)\*** | **(N=10)** |
| **Cardiac disorders** | 2 (9.09) | - |
| Pericardial effusion | 1 (4.55) | - |
| Sinus tachycardia | 1 (4.55) | - |
| **Endocrine disorders** | 1 (4.55) | - |
| Hypothyroidism | 1 (4.55) | - |
| **Gastrointestinal disorders** | 1 (4.55) | - |
| Constipation | 1 (4.55) | - |
| **General disorders and administration site conditions** | 5 (22.72) | - |
| Chills | 5 (22.72)** | - |
| **Immune system disorders** | 1 (4.55) | - |
| Anaphylactic reaction | 1 (4.55) ** | - |
| **Infections and infestations** | 1 (4.55) | 3 (30.00) |
| Fungal infection | - | 1 (10.00) |
| Lower respiratory tract infection | - | 2 (20.00) |
| Urinary tract infection | 1 (4.55) | - |

| **System Organ Class**<br>Preferred Term | **Arm A**<br>(N=22)* | **Arm B**<br>(N=10) |
| --- | --- | --- |
| **Injury, poisoning and procedural complications** | 1 (4.55) | - |
| Infusion related reaction | 1 (4.55)** | - |
| **Investigations** | 12 (54.55%) | 2 (20.00) |
| Alanine aminotransferase increased | 1 (4.55) | - |
| Fibrin D dimer increased | 1 (4.55) | - |
| Low density lipoprotein increased | 1 (4.55) | - |
| Lymphocyte count decreased | 11 (50.00)** | 2 (20.00) |
| Non-high-density lipoprotein cholesterol increased | 1 (4.55) | - |
| Platelet count decreased | 1 (4.55)** | - |
| **Metabolism and nutrition disorders** | 6 (27.27) | 1 (10.00) |
| Hyperglycemia | 4 (18.18) | 1 (10.00) |
| Hypertriglyceridemia | 2 (9.09) | 1 (10.00) |
| **Respiratory, thoracic, and mediastinal disorders** | - | 3 (30.00) |
| Acute respiratory distress syndrome | - | 2 (20.00) |
| Respiratory failure | - | 2 (20.00) |

\* 2 subjects could not complete even one dosing and were replaced as per protocol. They are part of safety population set till their discontinuation.
\*\* Related to the study drug

Five patients (2 patients in Arm A out of the patients who received complete infusion and 3 patients in Arm B) reported serious adverse events (SAEs) during the study. The SAEs reported in Arm A were anaphylactic reaction and pericardial effusion. Anaphylactic reaction resolved on the same day with medical intervention and was considered as related to the study drug infusion. Pericardial effusion was considered due to underlying hypothyroidism. The patient was treated with levothyroxine and recovered. The event was considered not related to the study drug.

Three deaths were reported in Arm B. The first death was due to lower respiratory tract infection with ARDS; the second was due to type 1 respiratory failure with ARDS, with lower respiratory tract infection; and the third was due to respiratory failure. No fatal TEAEs were reported in Arm A.

As defined in the protocol, patients who did not complete one full dose were considered unevaluable and were replaced. Two patients randomized to Arm A, experienced an infusion reaction shortly after initiation of drug and did not complete the first dose and withdrew from the study. The event of infusion reaction resolved on the same day in both patients. Subsequently one patient recovered from COVID-19 in approximately 2 weeks and was discharged from the hospital. The second patient developed further complications of COVID-19 related ARDS and died 9 days after discontinuation and the event was deemed not related to the study drug.

All the infusion reactions (5 events) were considered related to study medication and resolved within a few hours with symptomatic management. These infusion reactions occurred when they were given over 2 hours. However, the reaction abated when the infusion was given over 5-6 hours.

No notable differences were seen between the arms in vital parameters and clinical laboratory (hematology and biochemistry) evaluations except lymphocyte count decrease, which was seen in 11 (50%) patients in Arm A and 2 (20%) patients in Arm B. The events in Arm A, although severe in nature, were transient, without any clinical consequences, and considered to be related to the study drug. The events can be attributed to the mechanism of action of the drug and expected in keeping with the known safety profile of the drug.

## Discussion

Progression of COVID-19 is associated with systemic hyperinflammation and elevation of inflammatory markers. As a consequence of this exaggerated immune response, there is a pro-inflammatory cytokine release syndrome, which is associated with high mortality in COVID-19 patients [26]. In patients in intensive care, who are critically ill (require ventilation) and seriously ill (require oxygen support), there is an increased concentration of IL-2, IL-7, IL-10, GM-CSF, IP-10, MCP1, MIP1a and TNF-a [5]. This inflammatory pathophysiology of COVID-19 has encouraged research into the use of immunomodulating treatments, such as Itolizumab, in moderate to severe cases of COVID-19, which was approved seven years ago for use in psoriasis in India and has also been used in rheumatoid arthritis [27–29]. Further, the mechanism of action of Itolizumab ensures immunomodulating effect by acting upstream in the Th1 and Th17 pathways, thus providing additional benefits over other similar agents [30,31].

Building on the experience of using Itolizumab, its documented safety profile, and its mechanism of action, we undertook the current study to explore its potential to prevent cytokine release syndrome and reduce mortality in moderate to severe ARDS in COVID-19 patients. The current effort provides encouraging results, particularly with respect to mortality noted at the end of 30 days follow-up, and there is a need to replicate these findings either through additional, larger clinical trials or post-marketing surveillance studies.

With the global caseload of COVID-19 edging past 60 million, and the death tally having crossed a million, it is imperative to explore therapeutic alternatives which can not only prevent progression to severe disease, but also reduce mortality and morbidity before clinical response capacities are overwhelmed [1,32]. We identified a mortality benefit in the current study and interpret this cautiously considering the small sample size and usual limitations of undertaking an open label study, which has been conducted within the restrictions imposed by an ongoing pandemic [33]. We further acknowledge that the open-label design is also known to yield slightly larger estimates of effect size, to ameliorate which blinded, large clinical trials need to be conducted [34]. However, what is encouraging is that in addition to the mortality benefit, the current effort also identified favorable outcomes related to improved lung functions, biomarker profile and clinical resolution, especially with respect to respiratory/ventilatory support requirements. This constellation of clinical and laboratory findings supporting the beneficial effect of Itolizumab, which are likely to be internally valid for the given patient set, warrants deeper investigation to ensure generalizability and external validity of the results. The recent emergency use authorization accorded to Itolizumab for use in COVID-19 patients in India and Cuba, provides a window of opportunity to conduct a larger, global, phase 3 study and undertake post-marketing surveillance to explore the utility and impact of Itolizumab in COVID-19 cases with cytokine release syndrome.

The role of systemic vasculitis and cytokine mediated coagulation disorders have been recognized as significant factors for multi organ failure in patients with severe COVID-19 complications [35]. In patients with respiratory distress, levels of organ dysfunction markers such as D-dimer and lactate dehydrogenase (LDH) and surrogate markers of inflammation or cellular damage, such as ferritin and CRP, need to be closely monitored as they are considered as markers for potential progression to critical illness [36–38]. Elevated LDH levels indicate acute inflammation and have been associated with a 6-fold increase in the odds of progressing to severe COVID-19 and a 16-fold increase in odds of dying from COVID-19 [39]. D-dimer levels indicate coagulopathy and higher values have been shown to be associated with poorer clinical outcomes [40,41]. Serum ferritin levels indicate RBC damage and have been identified to be independently associated with the development of severe COVID-19 [42]. Plasma CRP levels have also been associated with CT confirmed moderate to severe pneumonia in COVID-19 patients [43]. Encouraging trends were noted for all these biomarkers in Arm A patients, who received Itolizumab in addition to best standard of care. A consistent reduction in D-dimer and LDH levels were seen in Arm A unlike in Arm B where no pattern was observed. The mean change in ferritin from baseline was higher in Arm A at all timepoints. Further, decreasing levels of these markers was accompanied by clinical improvement in patients receiving Itolizumab.

A four-fold decrease was seen in IL-6 levels in Arm A after the administration of Itolizumab (p=0.0269) while a 30% increase was observed in Arm B. Our findings are also in agreement with the preliminary findings from a small study from Cuba, where reduction in IL-6 levels was seen in COVID-19 patients treated with Itolizumab [44]. TNF-α also followed a similar trend (four-fold decrease) as IL6 in Arm A after administration of Itolizumab (p= 0.025). In contrast, a three-fold increase in TNF-α was seen in Arm B.

A decrease in lymphocyte count has been observed in COVID-19 patients [45]. Absolute lymphocyte count is considered as an important prognostic marker in COVID-19 infection [46]. In Arm A, a transient reduction in ALC was seen by Day 7, which was considered related to the study drug. However, the levels increased from Day 7 to Day 30 and was comparable to Arm B, and this did not have any adverse clinical outcomes in the patients.

A total of five serious TEAEs were reported in the study of which 3 deaths were reported in Arm B. Of the two serious TEAEs reported in Arm A, one (pericardial effusion) was related to underlying comorbidity (hypothyroidism) and was unrelated to the drug. The other was anaphylaxis due to infusion reaction, which is a known adverse effect of Itolizumab. The reported anaphylactic reaction was due to the shorter duration of infusion (2 hours). Other than the serious case of infusion reaction, non-serious treatment related infusion reactions were reported which abated with the extension of infusion period to 5-6 hours. The TEAEs such as infusion reactions and related events reported in the study were those expected for a monoclonal antibody (in treatment of psoriasis with the study drug, 10-15% of patients experienced infusion related reaction) [20]. The other treatment related AE was lymphocyte count decrease which was transient, and the patients recovered. In general, immunomodulatory drugs are expected to increase the risk of infection by acting on the immune system. However, in this study only one case of unrelated infection was reported in Arm A. These results are in line with the earlier finding with Itolizumab [17,19,22,29,47].

Considering the paucity of clinical therapeutic alternatives for COVID-19, the current effort highlights Itolizumab as a promising prospect deserving further study. Several ‘repurposed’, ‘emergency use’ or ‘off-label’ drugs are being considered as treatment alternatives for management of cases in the earlier phase of the disease, when interfering with viral replication may provide clinical benefits. Hydroxychloroquine has been shown to have limited clinical effectiveness [48–54], and Remdesivir [55,56] Favipiravir [57,58] and convalescent plasma [59–61] remain under investigation. If the patient enters the mechanical ventilation phase or when the patient’s condition is deteriorating despite oxygen administration, corticosteroids like methyl prednisolone and dexamethasone can be administered to prevent inflammation and further reduce mortality [35,62]. Other interventions like heparin to prevent blood clots and thrombogenic response, antibiotics such as azithromycin and ivermectin to reduce infections continue to be used in mild to moderate cases of COVID-19.

Immunomodulatory drugs find use in COVID-19 when the inflammatory cascades are starting to get activated. Tocilizumab, an IL-6 inhibitor, used in rheumatoid arthritis, has been repurposed for use in COVID-19, and has also received emergency use authorization in India. However, it remains limited by the fact that it blocks only IL-6, has no T-cell mediated immunomodulation, and has a short duration of action due to its downstream point of action [30,63–68]. Itolizumab has a broad immunological window being a CD6 inhibitor and is an option in the treatment of cytokine release syndrome in COVID-19 patients.

## Conclusion

The current investigation highlights the potential of Itolizumab as a promising, safe and effective immunomodulatory therapy for COVID-19 patients with cytokine release syndrome, as it efficiently controls immune hyperactivation, resulting in reduction in morbidity and mortality from moderate to severe COVID-19.

## Data Availability

All data reported in the manuscript are present in the company database.

## Acknowledgements

Editorial assistance was provided by Shivani Mittra, PhD and Ubhayabharathi Gurunath, M.Sc (Biocon Biologicals Ltd.).

Clinical support was provided by Arpitkumar Prajapati, M.D., Radhika A, M.D., and Sarika S Deodhar, M.D. (Biocon Biologicals Ltd).

Trial operation support was provided by Anirudh Sahoo, M.Pharm (Biocon Biologicals Ltd).

Sandeep N. Athalye, as the guarantor of this work, takes full responsibility for the work as a whole, including the study design, access to data, and the decision to submit and publish the manuscript.

## Authorship Statement

All authors were involved in the design of the clinical study, analyzed and interpreted the study data and results. All authors participated in the preparation and review of the manuscript. All authors read and approved the final version of the manuscript.

